# Convolutional neural networks on eye tracking trajectories classify patients with spatial neglect

**DOI:** 10.1101/2020.07.02.20143941

**Authors:** Benedetta Franceschiello, Tommaso Di Noto, Alexia Bourgeois, Micah M. Murray, Astrid Minier, Pierre Pouget, Jonas Richiardi, Paolo Bartolomeo, Fabio Anselmi

**Affiliations:** The LINE (Laboratory for Investigative Neurophysiology), Department of Diagnostic and Interventional Radiology, Lausanne University Hospital and University of Lausanne, Lausanne, Switzerland; Department of Ophthalmology, Fondation Asile des Aveugles and University of Lausanne, Lausanne, Switzerland; Laboratory of Cognitive Neurorehabilitation, Faculty of Medicine, University of Geneva, Geneva, Switzerland; CIBM Center for Biomedical Imaging, Lausanne, Switzerland; Department of Hearing and Speech Sciences, Vanderbilt University, Nashville, TN, USA; Sorbonne Université, Inserm, CNRS, Institut du Cerveau - Paris Brain Institute, ICM, Hôpital de la Pitié-Salpêtrière, Paris, France; Center for Neuroscience and Artificial Intelligence, Department of Neuroscience, Baylor College of Medicine, Houston, TX, USA; Center for Brains, Minds, and Machines, McGovern Institute for Brain Research at MIT, Cambridge, MA, USA; Department of Radiology, Lausanne University Hospital and University of Lausanne, Lausanne, Switzerland

**Keywords:** neglect, bio-markers, eye-tracking, deep networks, structural lesion, diffusion tensor imaging

## Abstract

**Background and Objective:** Eye-movement trajectories are rich behavioral data, providing a window on how the brain processes information. We address the challenge of characterizing signs of visuo-spatial neglect from saccadic eye trajectories recorded in brain-damaged patients with spatial neglect as well as in healthy controls during a visual search task.

**Methods:** We establish a standardized preprocessing pipeline adaptable to other task-based eye-tracker measurements. We use a deep convolutional network, a very successful type of neural network architecture in many computer vision applications, including medical diagnosis systems, to automatically analyze eye trajectories.

**Results:** Our algorithm can classify brain-damaged patients vs. healthy individuals with an accuracy of 86±5%. Moreover, the algorithm scores correlate with the degree of severity of neglect signs estimated with standardized paper-and-pencil test and with white matter tracts impairment via Diffusion Tensor Imaging (DTI). Interestingly, the latter showed a clear correlation with the third branch of the superior longitudinal fasciculus (SLF), especially damaged in neglect.

**Conclusions:** The study introduces a new classification method to analyze eyes trajectories in patients with neglect syndrome. The method can likely be applied to other types of neurological diseases opening to the possibility of new computer-aided, precise, sensitive and non-invasive diagnosing tools.

**Highlights:**

- We identify signs of visuo-spatial neglect through an automated analysis of saccadic eye trajectories using deep convolutional neural networks (CNNs).
- We provide a standardized pre-processing pipeline adaptable to other task-based eye-tracker measurements.
- Patient-wise, we benchmark the algorithm prediction with standardized paper-and-pencil test results.
- We evaluate white matter tracts by using Diffusion Tensor Imaging (DTI) and find a clear correlation with the microstructure of the third branch of the superior longitudinal fasciculus.
- Deep CNNs can efficiently and non-invasively characterize left spatial neglect.

## 1. Introduction

Eye-movements are non-invasive and readily accessible behavioral read-outs, providing a window onto how the brain processes information. The behavioral performance of the eyes, in particular via saccadic eye-movements, has been the focus of decades of research linking functional oculomotor behavior to dysfunction [2]. For instance, saccadic eye-movements can be a precursor of brain pathology and may also constitute an important bio-marker for early diagnosis of brain impairments [34]. They may also be particularly affected after a focal brain lesion, such as in patients suffering from neglect [37, 11, 38].

Left visuo-spatial neglect (hereafter simply ‘neglect’) is a frequent, but still poorly understood neurological syndrome that is characterized by a lack of awareness of contralesional stimuli following right hemispheric damage [6]. The diagnosis of neglect is important since this syndrome is associated with poor functional outcomes [28]. A high degree of overlap between attentional orienting deficits in neglect patients and their oculomotor performance has been demonstrated. Neglect patients exhibit saccadic impairments including direction-specific deficits of saccadic production [12, 48], saccadic amplitude or difficulty retaining locations across saccades [26]. Previous studies have shown that saccadic eye-movements are a sensitive measurement to characterise neglect [29]. This is an important observation since paper-and-pencil tests administered to evaluate neglect have limited sensitivity[3]. Furthermore, the neural mechanisms underlying neglect also still remain debated. Neglect has been linked with structural damage of key parietal regions such as the temporo-parietal junction (TPJ) or the inferior parietal lobule [20]. Other studies indicated that damage of long-range white matter tracts, connecting frontal and parietal areas, may represent crucial antecedent of neglect [8, 19]. Neglect has been reported following damage to the second and third branches of the superior longitudinal fasciculus (SLF) or to the inferior fronto-occipital fasciculus (IFOF), disrupting connectivity within attentional network of the brain [44, 43].

Convolutional neural networks (CNNs) have proven to be extremely successful algorithms, particularly in image classification [33], where they can outperform human observers and experts on some tasks. CNNs are versatile function approximators capable of detecting subtle patterns that are crucial for the final task (e.g. classification). Thanks to the enormous amount of data generated within the healthcare sector, recently CNNs have been successfully employed in a variety of image-driven medical diagnosis domains (e.g. radiology [18, 49], ophthalmology [39]). Current research on machine learning techniques applied to eye-tracking data have hitherto focused on different fields, such as classification of eye movements (fixation, saccades, etc.) [50], or computer-assisted diagnosis tools [30]. Other machine learning techniques provide early diagnosis methods for classification and detection of neuro-developmental disorders [41], methods aiming at detecting the presence of strabismus [17], or deep learning methods for the detection of Alzheimer’s disease [14, 13], or Mild Cognitive Impairment [32] based on eye-movement behavior identification. Among the latter, in [14], the authors use deep neural networks to identify patients with Alzheimer disease (AD). The study highlights how in principle CNNs can be used early diagnosis of AD. As for neuro-developmental disorders [40] and strabismus [17], CNNs were employed to capture subtle geometric features of eye trajectories, to which common diagnostics could be blinded to, to inform about the patient’s status. A support vector machine algorithm successfully classified patients with memory impairments [32], or readers with dyslexia [41] to classify readers with dyslexia.

The contributions of the present work are threefold. First, the pre-processing of eye-tracker trajectories is often not detailed in its steps, despite data needing to undergo cleaning and re-organisation prior to analysis. Eye-tracking data typically contain errors and noise that must be accounted for [25]. We provide an overview and code of the pre-processing required, and standardized it, to lay out a pipeline adaptable to other task-based eye-tracker measurements. Second, this paper demonstrates how modern machine learning algorithms, in particular CNNs, allow learning of representations of data features that are particularly effective in classifying pathological versus non-pathological conditions from patterns of eye-movements. Our method stands within the growing field of automatic diagnostic tools, a branch of non-invasive techniques at the interface between neuroscience and computer science, which can transform a simple task into an automatic diagnostic procedure. Finally, we explored the anatomical correlates of eye-tracking trajectories at a network level, by using diffusion tensor imaging (DTI). To the best of our knowledge, this is the first time that CNNs are used to determine and quantify the presence of neglect through eye-movement analysis during a visual search task and that a link between the algorithm outputs and anatomical markers is established.

## 2. Material and Methods

### 2.1. Behavioral and neuroimaging data collection

#### Participants

We analyzed eye movement data in a sample of 15 right-brain damaged patients with left visuo-spatial neglect and in 9 healthy controls, recruited from a previous study [15] Seven of the patients had varying degrees of a left visual field defect, as assessed by confrontation testing or perimetry. Demographic and clinical characteristics of the patients are presented in Table 1 (1, A). Healthy individuals were age-matched with the patients (mean age 58 years, range 45-69, *t* < 1) and had no neurological or psychiatric history.

**Table 1:** Classification results across the 10 random runs using either the x or the y spatial coordinates. Values are presented as mean ± standard deviation. Bold values indicate the highest performances. Coord = spatial coordinate; Acc = accuracy; Sens = sensitivity; Spec = specificity; PPV = positive predictive value; NPV = negative predictive value; F1 = F1-score; AUC = area under the ROC curve; AUPR = area under the PR curve.

| Coord | Acc (%) | Sens (%) | Spec (%) | PPV (%) | NPV (%) | F1 (%) | AUC | AUPR |
| --- | --- | --- | --- | --- | --- | --- | --- | --- |
| **x** | 86 ± 5 | 80 ± 11 | 89 ± 3 | 82 ± 6 | 88 ± 6 | 81 ± 8 | .85 ± 0.06 | .85 ± 0.06 |
| **y** | 85 ± 3 | 79 ± 6 | 88 ± 4 | 80 ± 5 | 88 ± 3 | 79 ± 4 | .83 ± 0.03 | .83 ± 0.03 |

#### Apparatus, stimuli and procedure

Participants were asked to perform a visual search task (Fig. 1) [15]. Participants saw 8 circles around a central circle and had to search for a peripheral circle missing its upper or lower part (target). Participants were required to maintain their gaze on the central circle and to freely move their eyes as soon as the central circle disappeared. Eye-movements were recorded at a sampling rate of 300Hz with a Tobii TX300 eye-tracker. The cue lasted for 3000ms. On each trial, one quadrant of the central circle was highlighted in white, serving as a predictive attentional cue, and indicating the most likely location of target appearance. The cue correctly indicated the target location on 73% of the trials (valid location). The target appeared in one of the three uncued quadrants (invalid location) on 18% of the trials. The target was not present on the remaining 9% of the trials (catch trials), which were included in the design to avoid guesses and anticipations. The target was then presented after a random interval ranging from 1000 to 2000ms. The target was created by deleting either the upper or the lower part (0.4^?^ of visual angle) of one of the eight peripheral circles. The color of each target and distracters changed randomly in each trial, thus requiring an attention-demanding serial search. Participants were asked to move a joystick up when the upper part of the circle was missing, or down when the lower part was missing, as fast and as accurately as possible with their right hand. The target disappeared when a response occurred, or after 3000ms if no response was made. This experiment was composed of a total of 176 trials.

**Figure 1:**
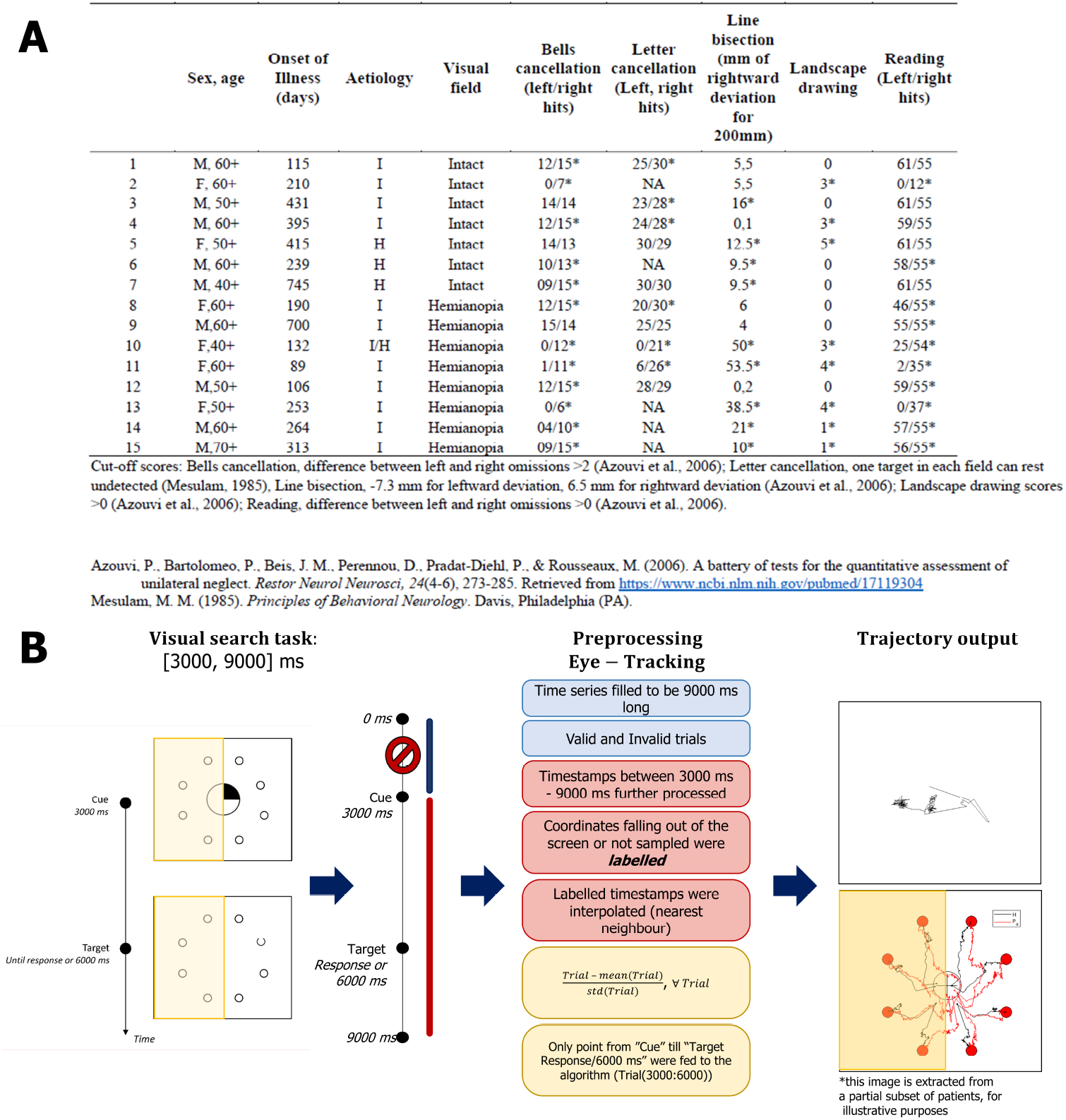
(Top). A. Demographic and clinical characteristics of neglect patients, with their performance on visuo-spatial tests. Asterisks denote pathological performance compared to normative data. For line bisection, positive values indicate rightward deviations, negative values indicate leftward deviations. Scores for the landscape drawing [22] indicate the number of omitted left-sided details. I, ischemic; H, hemorrhagic; NA, not available. B. The preprocessing steps required to analyse the eye-tracker data and the corresponding parts of the visual search behavioral paradigm task they refer to. (Bottom-right) An example of pre-processed trial, as well as the mean trajectories over all trials of a subset of subjects for each target. In the bottom row the left yellow box refers to the left-sided targets used for the analysis.

#### Neuroimaging data collection

Diffusion Tensor Imaging (DTI) tractography was used to study long-range of sub-cortical white matter pathways. For the complete preprocessing pipeline, see [15]. The mean fractional anisotropy values of the three branches of the superior longitudinal fasciculus (SLF), the cingulum, and the inferior fronto-occipital fasciculus (IFOF) were extracted in the right hemisphere. These tracts were chosen on the basis of their implication in attention networks and in visual neglect [8]. The analysis was conducted on n=13 patients, because the MRI scans were not available for two patients.

### 2.2. Data processing and analysis

#### Preprocessing

Targets presented within the left visual field were considered for the analysis, as neglect symptoms concern first and foremost attentional orienting towards the left, contralesional hemispace. Recorded eye-tracking trajectories underwent a pre-processing procedure to standardize the dataset before analyzing it. The code was developed with Matlab R2019b [36]. First, the duration of the trajectories was standardized across trials, i.e. all trials were uniformed in order to be 9s long (9000 ms). Time series corresponding to trials ending before 9*s* were filled with NaN (not a number, missed recording). The analysis focused on those trials both valid, invalid and not missed by the subject, according to the definition given in section 2.1. Only time-stamps recorded after the first 3000ms and before 9000ms were first taken into account. A loop over the total number of targets seen by each subject was iterated to analyze all trajectories recorded per each participant. For each target *i* the set of trajectories’ coordinates (*Eye*_*x*_, *Eye*_*y*_)*i* was extracted and coordinates corresponding to outliers (such as points outside the screen or above 9000ms) were respectively filled with “NaN” if acquired between 3000 and 9000ms, otherwise removed. All missed recordings were then re-filled according to the following criteria: if a NaN was present at the beginning of the trial, the NaN value was replaced with the center of the screen coordinates, at pixels (384, 512). If a NaN was in the middle of the time series, this value was interpolated (nearest neighbor) using the available neighboring sampled data, corresponding to actual recordings. If the target was reached at a certain timestamp, all remaining points (to reach the 9000ms upper limit) were filled with the target coordinates. The first 3000ms of *Eye*_*x*_ and *Eye*_*y*_ was erased, as they corresponded to the fixation part, leaving us with a shortened time series 6000ms long, now cleaned-up and interpolated. Also, the latter 3000ms were eliminated to avoid piece-wise constant trajectories, as participants tended to reach the target beforehand (see Figure 1). The post-processed time-series corresponds to the 3000ms of the visual search task. Every trial for every subject and every target *i*, (*Eye*_*x*_, *Eye*_*y*_)*i*, was z-scored by subtracting its mean and dividing by its standard deviation. All pre-processing steps are schematized in Figure 1. Only target presented to the left visual field of the participant have been considered.

#### Classifying healthy vs. neglect patients from eyes tracking images

In this section we discuss the methodology we applied to estimate healthy versus neglect patients’ status from a subset of eye-tracking trajectories, i.e. those corresponding to targets belonging to the left hemispace. Left hemispace targets are more challenging for patients affected by this syndrome. To this purpose, we formulated the estimation problem as a classification task, i.e. learning the mapping between an appropriate representation of the eyes’ trajectories and the patient’s label. In particular, we separately analyzed the *x* coordinates and *y* coordinates of the trajectories (*x*-projection and *y*-projection), of target presented to the left visual field of the participant. The resulting vectors have length *d* = 1001 and are used as input samples to our network.

We designed a custom CNN with building blocks inspired by the VGG-16 model [45]. Specifically, our network is composed of a sequence of 3 blocks of convolutional layers, each one separated by a pooling layer that halves the output vector dimension. A kernel size of 3 and zero padding were used in all convolutional layers. To obtain the desired classification output, the convolutional blocks are followed by three dense, fully connected (FC) layers. We applied the ReLU activation function for all layers, except for the last layer which is followed by a sigmoid function. We used Xavier initialization [24] for all layers. Biases were initialized to 0 and a batch size of 16 was chosen. Batch normalization [27] and dropout (rate = 0.3) [46] were used to avoid overfitting in the convolutional blocks and in the FC layers, respectively. To fit the model, the Adam optimization algorithm [31] was applied, with a learning rate of 0.0001. We trained the model for 25 epochs and we adopted the binary cross-entropy loss function. The total number of trainable parameters in our network is 264, 377. Training and inference were implemented using Tensorflow (version 2.5) [1]. The detailed structure of our network is illustrated in Figure 2. To compute unbiased test results, we applied a 5-fold cross-validation (CV) at the individual participant level. This ensured that multiple trajectories of the same participant did not appear both in the test and training set. Furthermore, to account for the variability introduced by the random choice of patients at each CV split, the whole CV was repeated 10 times (10 runs), each time performing the splitting anew, and results were averaged.

**Figure 2:**
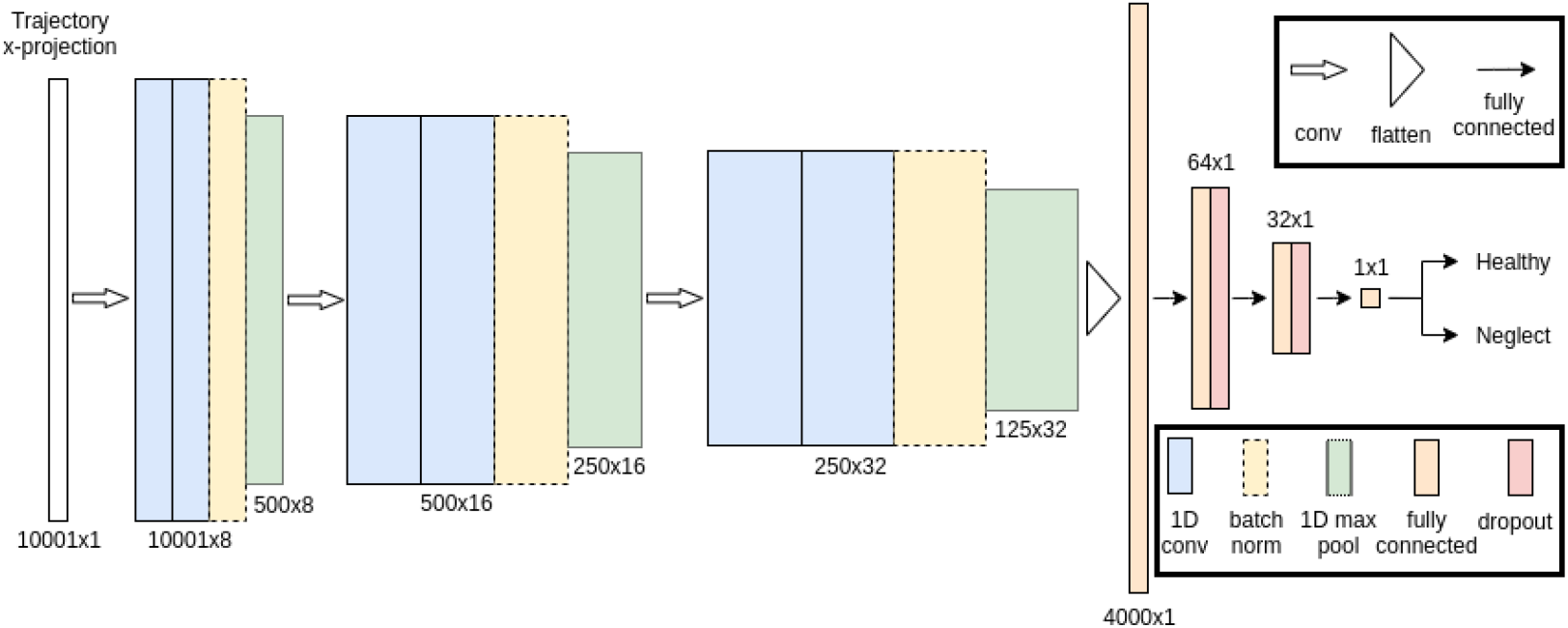
(Left). The CNN architecture used in our experiments. Three convolutional blocks with batch normalization and pooling are followed by two dense layers with dropout. Dimensions of each layer are reported in the image. The final output is the probability that each input trajectory has to either belonging to a healthy control or to a patient with spatial neglect. To assign the final class (healthy vs. neglect) to one subject, we performed a majority voting across all trajectories of that subject.

**Figure 3:**
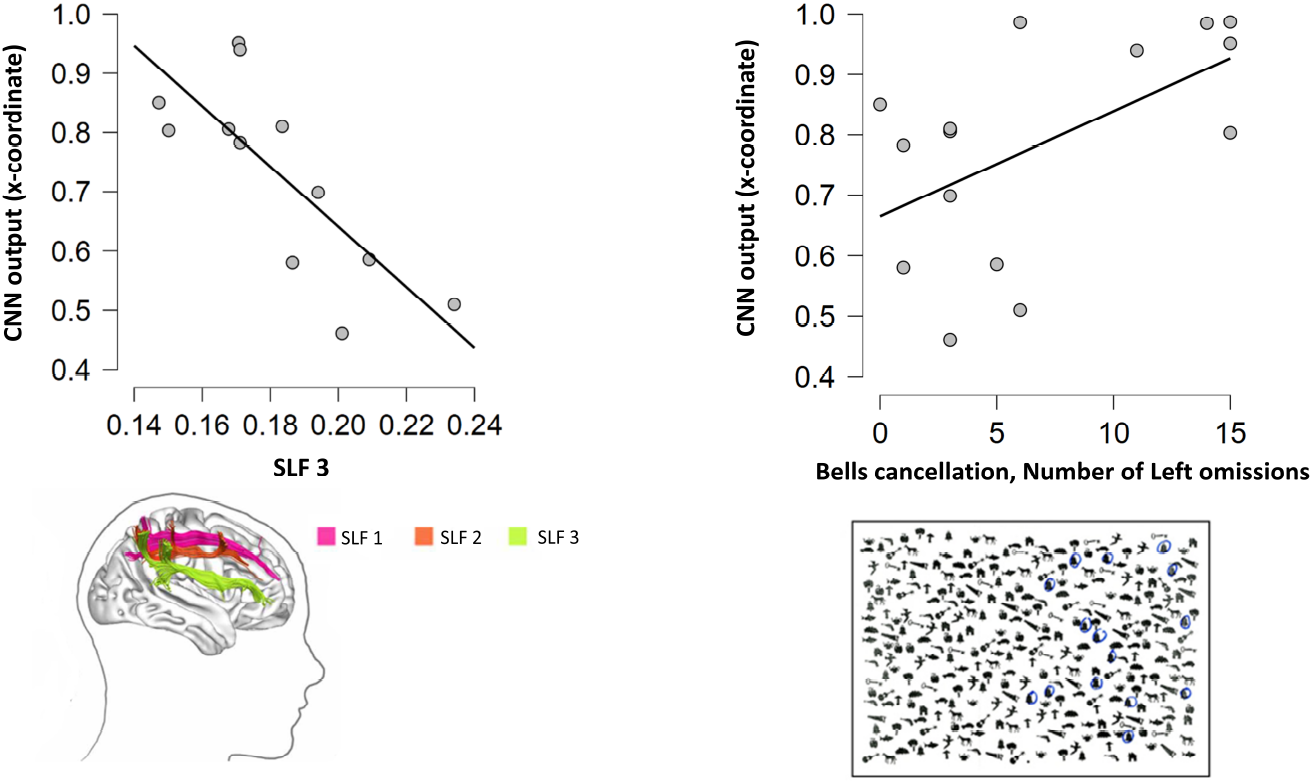
The correlation between the algorithm confidence scores and the Fractional Anisotropy (FA) of the SLF3 is presented (left panel) (*ρ* = −0.77), as well as the correlation between the algorithm confidence scores and the number of left omissions of the Bells cancellation test (right panel) (*ρ* = 0.55, *p* = 0.033). The images of depicting the SLF and the Bells test are reprinted from [10].

Finally, we computed the classification accuracy for each participant. This corresponds to the ratio between the correctly classified trajectories and the total number of trajectories. It can be interpreted as a measure of confidence *c* with respect to the final prediction. For instance, *c* = .99 implies an extremely confident correct classification; *c* = .65 implies a moderately confident correct classification; *c* < .5 implies a wrong classification. To obtain one single confidence score per participant, we averaged the confidence scores of each participant across the 10 random runs.

The developed code can be adapted to the analysis of visual search tasks and possibly integrated in existing algorithms, such as those identifying fixation and regions of interest of eye-tracker trajectories [16] as well as those assessing data quality [42].

## 3. Results

### Behavioral results

In [15], oculomotor behaviour was studied together with manual responses on a visual search task in neglect to determine the relation between saccadic parameters and sub-clinical disorders of spatial attention. The study showed the occurrence of inappropriate rightward saccades during target selection: when left-sided targets were presented, saccades were equally likely to be performed towards the left side or towards the right hemispace. Right-sided distracters may erroneously capture patients’ attention, leading to an over-exploration of the right hemispace, consistent with the magnetic attraction of gaze typically observed in neglect [21]. Thus, pathological production of eye movements should be considered as a subtle manifestation of visuo-spatial disorders.

### Classification results

Table 1 illustrates the classification test results averaged across the 10 random runs when using the *x* and the *y* projections. Overall, the network performances are competitive regardless of the spatial coordinate utilized with a slightly better performance for the *x* projection, in agreement with the observations in [37]. We focused on a 1-dimensional analysis to reduce the computational time of the pipeline and to simplify the model, which is convenient for studying its integration in the clinical practise.

The results we obtained are in line with both a technical and clinical perspective. On the one hand, as mentioned in section 1, patients’ healthy vs. unhealthy status can indeed be evaluated extracting salient features from the geometry of eye-trajectories in visual tasks, using a deep convolutional neural network. On the other hand, neglect features can be equally identified either from *x* or *y* trajectories [37], although the performance is more efficient on *x*-ones.

#### CNN results and Neuroimaging

We performed Pearson correlations between the algorithm confidence score on the *x* and *y*-spatial coordinate and the Fractional Anisotropy (FA) of long-range of white matter tracts connecting attentional networks. One patient was considered as an outlier and was discarded from the analysis. Our results show a negative correlation (*ρ* = −0.77) between the algorithm confidence score on the *x*-spatial coordinate and damage of the SLF3 (*p* = 0.003 Bonferroni-corrected, see 3, left panel). The correlations with the others tracts (SLF1, SLF2, IFOF, Cingulum), as well as between the *y*-coordinate and all the other tracts did not reach significance (all Bonferroni- corrected *p >* 0.011). Also, the algorithm score on the *x*-spatial coordinate correlated with the number of left omissions in Bells cancellation [23], a standardized paper-and-pencil test use to diagnose neglect signs (*ρ* = 0.55, *p* = 0.033) 3, right panel).

## 4. Discussion and conclusions

Neglect is a multi-component syndrome; dissociations in performance on different tests, are often observed, both between and within patients. Some of these dissociations may depend on the activity of compensatory mechanisms, such as top-down orienting of attention [9], perhaps driven in part by the healthy left hemisphere [7]. Sensitive behavioral techniques, such as manual response times or eye movements characterisation, [4, 5, 37, 11, 38] may thus greatly help diagnosis of neglect.

Our work demonstrates how deep convolutional neural networks allow learning of representations of saccadic eye-movements’ features that are particularly effective in classifying pathological neglect versus non-pathological conditions with a reached accuracy of 86 ± 5%. To the best of our knowledge, this is the first time that CNNs are used to determine the presence of neglect through eye-movement analysis during a visual search task. The correlation between the algorithm confidence score output and the anatomical markers of the patients’ DTI benchmark the relevance of the technique and its specificity in detecting neglect patients. Furthermore, the algorithm confidence score on the *x*-coordinates of the saccades trajectories appears to be related to the impairment of the third branch of the superior longitudinal fasciculus (SLF3), *ρ* = −0.77, *p* = 0.003. The SLF3 links parietal and frontal regions and has been shown to be specifically impaired in neglect [35, 47]. We also observe a correlation between the algorithm confidence score on the *x*-coordinates and the degree of severity of neglect signs (*ρ* = 0.55, *p* = 0.033). Although the algorithm confidence scores are similar for the *x* and *y*-projections of the saccades, the correlations with the anatomical and behavioral data show that predominant features seem to be contained in the *x*-projections.

Despite several studies highlighted pathological eye-movement behavior as a consequence of impaired shift of attention in neglect, the measurement of eye-movement behavior as a tool to diagnose neurological pathology has only begun to be studied. In this respect our work represents therefore a contribution to understand and predict the trajectory of individual patients, opening to the possibility of a new computer-aided diagnosis tool for neglect syndrome. Also, the classification could allow us to assess the level of impairment and assist follow-up treatment. Further investigations should consolidate this link, allowing to differentiate and predict patterns in agreement with the anatomical markers with unprecedented precision, especially as the neural substrates of neglect are still debated, despite this syndrome representing a unique opportunity to underpin the underlying mechanisms of spatial processing and conscious awareness. Finally, this work represents a first step towards the scalability of deep learning techniques to other neurological conditions characterised by impaired eye movements: a growing and exciting field of investigation.

## Data Availability

The dataset is detailed, analysed and discussed in https://www.sciencedirect.com/science/article/abs/pii/S0028393215001591
In the current work we only used the dateset for supplementary automatized analyses.

## 5. Acknowledgments

B. F. and A. M acknowledge the support of the Fondation Asile des Aveugles. M. M. M. acknowledges the support of grants from the Swiss National Science Foundation (grant 320030-169206), the Fondation Asile des Aveugles, and a grantor advised by Carigest SA. B. F and M. M. M. acknowledge access to the facilities and expertise of the CIBM Center for Biomedical Imaging, a Swiss research center of excellence founded and supported by Lausanne University Hospital (CHUV), University of Lausanne (UNIL), Écôle polytechnique fédérale de Lausanne (EPFL), University of Geneva (UNIGE) and Geneva University Hospitals (HUG). The work of P.B. is supported by the Agence Nationale de la Recherche through ANR-16-CE37-0005 and ANR-10-IAIHU-06, and by the Fondation pour la Recherche sur les AVC through FR-AVC-017. F. A. acknowledges the Center for Brains, Minds and Machines (CBMM), funded by NSF STC award CCF-1231216 and the financial support of the AFOSR projects FA9550-17-1-0390 and BAA-AFRL-AFOSR-2016-0007 (European Office of Aerospace Research and Development), and the EU H2020-MSCA-RISE project NoMADS - DLV-777826.

